# Making colonoscopy-based screening more efficient: a ‘gateopener’ approach

**DOI:** 10.1101/2021.09.14.21263600

**Authors:** Thomas Heisser, Rafael Cardoso, Tobias Niedermaier, Michael Hoffmeister, Hermann Brenner

## Abstract

**Objectives:** To assess the potential of an innovative approach to colonoscopy-based screening for colorectal cancer (CRC), by use of a single, low threshold fecal immunochemical test (FIT) as a ‘gateopener’ for screening colonoscopy.

**Design:** Simulation study using COSIMO, a validated Markov-based simulation model, in a hypothetical German population.

**Setting:** Modelled scenarios included either direct invitation to screening colonoscopy or mailing a single (‘gateopener’) FIT along with an invitation to colonoscopy contingent on a FIT value above a low threshold yielding a 50% positivity rate (i.e., every other pre-test will be positive). The main analyses focused on scenarios assuming identical colonoscopy uptake, resulting from higher adherence to the gateopener FIT than to primary use of colonoscopy and avoidance of colonoscopy in those with below-threshold FIT values.

**Participants:** Hypothetical cohorts of 100,000 previously unscreened men and women using screening at different ages and with varying levels of adherence.

**Interventions:** Screening colonoscopy without and with preceding gateopener FIT.

**Main outcome measure:** Detected and prevented CRC cases and deaths within 10 years.

**Results:** Across all ages and both sexes, use of screening colonoscopy contingent on a positive gateopener FIT yielded approximately doubled cancer detection rates as compared to conventional screening. In those spared from undergoing screening colonoscopy due to a negative FIT pretest, numbers needed to screen were 10-times higher as compared to those for individuals with a positive FIT, peaking in more than 2500 and more than 3800 (hypothetically) needed colonoscopies to detect one case of cancer in 50-year-old men and women, respectively. At identical levels of colonoscopy use, gateopener screening resulted in 51-53% and 63-68% more prevented CRC cases and deaths, respectively.

**Conclusions:** By directing colonoscopy capacities to those most likely to benefit from it, offer of screening colonoscopy contingent on a ‘gateopener’ low-threshold FIT would substantially enhance efficiency of colonoscopy screening.

**Summary Box:** *What is already known on this topic:*

- Screening colonoscopy as primary examination is inefficient as most of the screened subjects would never develop colorectal cancer even without screening.
- Efficiency could be enhanced by pre-selecting those most likely to benefit, e.g., by use of a single low-threshold faecal immunochemical test (‘gateopener’ FIT)

*What this study adds:*

- A simulation where only individuals with positive gateopener FIT proceeded to screening colonoscopy resulted in 50% fewer colonoscopies required to detect one case of cancer vs conventional screening colonoscopy.
- At identical colonoscopy uptake rates, the gateopener approach implied approximately 50% and 70% more prevented colorectal cancer cases and deaths, respectively.
- Inviting subjects to undergo pre-testing with low-threshold FITs would markedly improve efficiency of colonoscopy-based screening.

## Introduction

Screening colonoscopy is the most sensitive method to detect early-stage tumors and CRC precursor lesions [1]. However, adherence to this invasive screening procedure is much lower than for less invasive primary screening offers [2–5], colonoscopy capacities are limited in many countries [6], and the majority of screening colonoscopy participants do not benefit from the exam as no clinically relevant neoplasms are detected and removed [7].

In this study, we present a novel approach to colonoscopy-based screening by use of a single, low threshold fecal immunochemical test (FIT) as a ‘gateopener’ for screening colonoscopy. In this approach, rather than inviting all eligible individuals directly to screening colonoscopy, invitation letters to undergo screening would include a FIT with very low hemoglobin detection thresholds, e.g., adjusted to yield a 50% positivity rate. Participants would be asked to send the FIT back to a central laboratory, and only those above the threshold would be invited to follow-up with screening colonoscopy.

The gateopener approach could be designed in such a way that the overall colonoscopy uptake rates would remain the same as for offers of colonoscopy as primary screening exam, as a result of higher adherence to gateopener FIT on the one hand, and selective colonoscopy uptake after the gateopener FIT on the other hand. The rationale for such a gateopener model would be that, at comparable colonoscopy uptake rates, much higher diagnostic yield could be achieved in comparison with an exclusive offer of screening colonoscopy as primary exam, and available screening colonoscopy capacities would be used much more efficiently by focusing on those who are most likely to benefit from this more invasive exam. We set up a modelling study to evaluate such a gateopener screening approach in terms of detection rates of CRC and its precursors, and reductions of CRC incidence and mortality when compared to conventional screening colonoscopy.

## Methods

### Multistate Markov Model

For this study, we used the previously developed and validated Markov-based <u>Co</u>lorectal Cancer Multistate <u>Si</u>mulation <u>Mo</u>del (COSIMO) to simulate effects of screening for CRC in a hypothetical German population [8]. Briefly, COSIMO simulates the natural history of CRC based on the process of precursor lesions developing into preclinical and then clinical cancer in a hypothetical population for a predefined number of years. Screening can interfere with the natural history of CRC (**Figure 1**).

**Figure 1.**
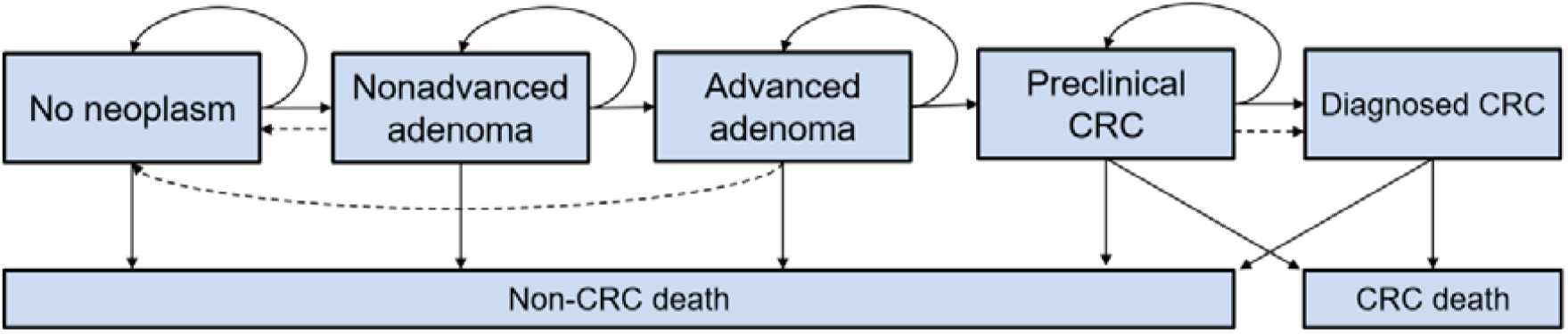
Schematic illustration of the <u>Co</u>lorectal Cancer Multistate <u>Si</u>mulation <u>Mo</u>del (COSIMO) Solid lines represent the progression of colorectal disease through the adenoma-carcinoma sequence in the absence of screening; dashed lines show the movement between states because of the detection and removal of adenomas and the detection of asymptomatic CRC at screening. CRC: Colorectal cancer.

The model’s natural history assumptions were derived step-by-step in several previous analyses using data from the German screening colonoscopy registry, the world’s largest registry of its kind [9–11]. Death rates from CRC were estimated using data from a large population-based case-control study with long-term follow-up of CRC cases and registry data from Germany as previously described [12,13]. General mortality rates and average life expectancy were extracted from German population life tables [14]. A comprehensive documentation on the model’s structure and data sources used for its development is given in **Supplementary Appendix 1**. Overviews of key model parameters are provided in **Supplementary Tables 1-3**. The model source code, developed in the statistical software R (version 4.0.2), is available for download from our website [15].

### Diagnostic Performance Parameters

For the gateopener FIT, we derived the sensitivity (the proportion of detected cases among all subjects with any adenomas or cancer) and specificity (the proportion of all subjects without adenomas or cancer correctly classified as such) of SENTiFIT-FOB Gold (Sentinel Diagnostics, Milan, Italy), a commonly used FIT test, adjusted for a predefined positivity level of 50%, i.e. using a positivity threshold that yields 5-10 times more positive tests compared to using the FIT with the conventional threshold (**Supplementary Table 1)** [16,17]. Compared to conventionally used cut-offs, adjusting for a positivity rate of 50% results in substantially increased sensitivity to the account of significantly decreased specificity.

Analyses to derive diagnostic performance parameters were conducted based on a study among 1667 participants of screening colonoscopy in Germany from 2005 through 2010 who had provided pre-colonoscopy stool samples for quantitative FITs as previously described [17]. To increase robustness of estimates for the sensitivity of CRC, additional pre-treatment stool samples were analysed for 444 subjects with confirmed CRC diagnosis recruited since 2013. Details on populations, sample and data collection, specimen collection and handling, as well as statistical analysis have been reported elsewhere [17,18].

### Simulations

#### Modelled Scenarios

To explore the effect of gateopener screening, we simulated a range of scenarios in hypothetical populations consisting of each 100,000 previously unscreened men or women. Simulated subjects were assumed to be invited to undergo screening colonoscopy at varying ages (50, 55, 60, 65, or 70), either without or with gateopener FIT screening. Sex-specific baseline neoplasm prevalences for each age of screening were extracted from a previous analysis of more than 4.4 million screening colonoscopies in the German-screening eligible population [21].

To reflect uptake patterns in different real-life settings [2–5], we assumed varying levels of adherence between modelled scenarios (**Table 1)**. For invitations to conventional screening colonoscopy, these implied 10%, 20%, and 30% colonoscopy use for low, moderate, and high levels of screening uptake, respectively.

**Table 1.** Assessed scenarios of use of colonoscopy as a primary screening offer without and with gateopener FIT.

| Scenario | Parameter | Adherence Pattern |  |  |
| --- | --- | --- | --- | --- |
|  |  | Low | Medium | High |
| Invitation to conventional screening colonoscopy | Use of screening colonoscopy | 10% | 20% | 30% |
| Invitation to screening colonoscopy with gateopener FIT | Use of gateopener FIT | 25% | 50% | 75% |
|  | Positive result of gateopener FIT with 50% positivity rate | 12.5% | 25% | 37.5% |
|  | Use of colonoscopy after positive FIT | 10% | 20% | 30% |

FIT, fecal immunochemical test.

Corresponding analyses for use of a gateopener FIT assumed higher levels of initial FIT uptake (low: 25%, moderate: 50%, high: 75%), which is in line with higher utilization levels for FIT vs endoscopic screening in practice [3–5]. As outlined above, gateopener FITs were assumed to yield a 50% positivity rate, i.e., every other test will be positive. Thus, low, moderate, and high uptake of the gateopener FIT would result in that 12.5%, 25%, and 37.5% of subjects, respectively, would have a positive result and be subsequently invited to undergo screening colonoscopy. Across scenarios, we assumed that 80% of those invited would make use of this colonoscopy offer, which reflects the observed compliance to colonoscopy after a positive test in real-world FIT-based screening [19]. Combined, these assumptions implied identical levels of total colonoscopy use as for conventional screening colonoscopy, i.e., 10%, 20%, and 30% for low, moderate, and high levels of initial screening uptake, respectively.

It should be noted that the gateopener approach can be adjusted to situations with different uptake rates and different gateopener FIT positivity thresholds. In order to facilitate comparability with respect to efficiency of colonoscopy use, we deliberately focused the primary analyses on scenarios yielding identical final colonoscopy uptake rates under plausible assumptions.

However, evidence on measures to ensure high levels of uptake to fecal testing [20], as well as on the generally high uptake of diagnostic colonoscopies [19], suggests that the design features of the proposed gateopener screening may likely result in higher final colonoscopy uptake rates. To cover this potentially more realistic outcome, we therefore also provide results for analyses for gateopener screening scenarios resulting in 50% higher final colonoscopy uptake rates (30% rather than 20%).

Finally, to explore the impact of uncertainty related to model key parameters, all point estimates of the transition rates between the various states in CRC development and progression were replaced by either the lower or upper limits of the 95% confidence intervals (CIs).

#### Outcomes

For each scenario, we firstly determined the expected detection rates for any advanced neoplasm (advanced adenoma and cancer) or cancer at all assessed screening ages by dividing the numbers of detected advanced neoplasms and cancers by the number of screening colonoscopy participants. In addition, to better contrast the performance of the gateopener approach, we derived the (hypothetical) colonoscopy detection rates for the same outcomes in those subjects who had a negative FIT result and who were thus spared from undergoing screening colonoscopy.

For all detection rates, we likewise report the reciprocal values, i.e., the numbers of colonoscopies needed to detect one advanced neoplasm or cancer for screening colonoscopies with and without gateopener FIT (NNS, number needed to scope). Advanced adenomas were defined as adenomas with at least 1 of the following features: ≥ 1 cm in size, tubulovillous or villous components, or high-grade dysplasia.

Furthermore, we determined the expected cumulative numbers of CRC cases and deaths within 10 years after screening and calculated and compared the corresponding reductions for gateopener screening scenarios to those for conventional screening scenarios. All analyses were carried out separately for men and women.

### Patient and Public Involvement

Patients and the public were neither involved in the design and conduct of this study, nor in writing or editing of this document. Research at the German Cancer Research Center (DKFZ) is generally informed by a Patient Advisory Committee.

## Results

Detection rates of colorectal cancer and of any advanced colorectal neoplasm are shown by sex and age in **Figure 2** and **Supplementary Figure 1**, respectively. They were generally higher among men than among women and increased with age. With conventional screening colonoscopy, cancer detection rates ranged from 0.2% among previously unscreened 50-year-old women to 1.9% among previously unscreened 70-year-old men (**Figure 2)**. While all of these cancer detection rates would be expected to almost double when screening colonoscopy was offered to a population preselected by a positive gateopener FIT, the (hypothetical) cancer detection rates in those spared from undergoing screening colonoscopy due to a negative FIT pretest were consistently very low (≤ 0.2%) across both sexes and all simulated ages.

**Figure 2.**
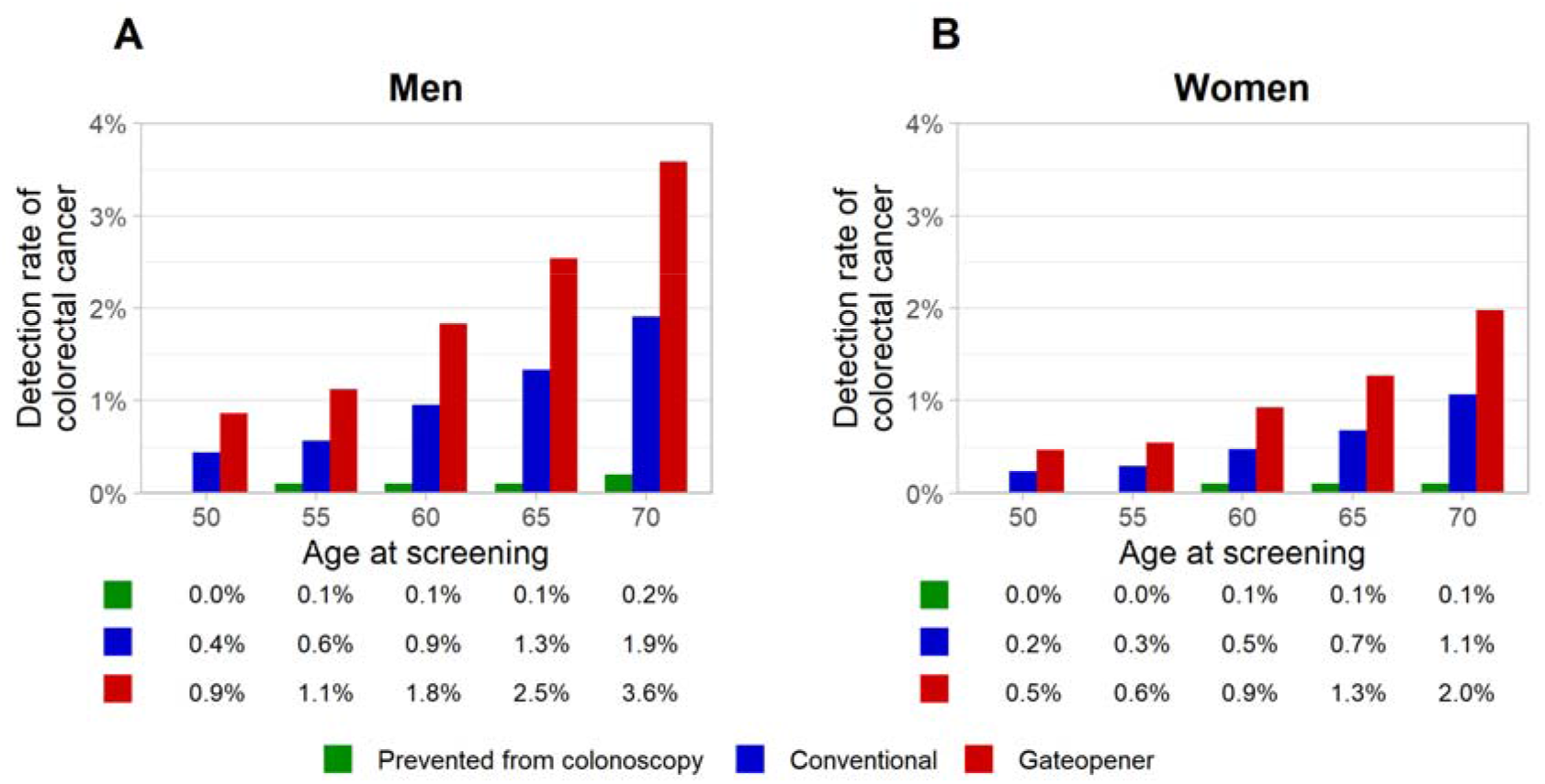
Detection rates of colorectal cancer at varying ages of colonoscopy screening without and with positive preceding gateopener FIT, and in those in fact prevented from undergoing screening colonoscopy due to a negative preceding gateopener FIT. (A) men; (B) women. CRC: Colorectal cancer, FIT: fecal immunochemical test

Similarly, detection rates of any advanced colorectal neoplasm, which ranged from 3% among previously unscreened 50-year-old women to 12% among previously unscreened 70-year-old men with conventional colonoscopy would be expected to increase by almost 50% using the gateopener approach, and approximately halve in those spared from undergoing screening colonoscopy (**Supplementary Figure 1)**.

Correspondingly, for both sexes and at all simulated ages, the NNS to detect one cancer or one advanced neoplasm was approximately halved or reduced by one third, respectively, as compared to conventional colonoscopy screening without preceding gateopener FIT **(Figure 3, Supplementary Figures 2)**. At age 50, a widely recommended age to start screening, pre-testing by a gateopener FIT was associated with approximately 100 and 200 fewer colonoscopies needed to detect one case of cancer in men and women, respectively. Notably, were screening colonoscopies to be conducted despite a negative gateopener FIT, approximately 10-times as many colonoscopies would be needed to detect a single case of cancer, reflecting the very low (hypothetical) detection rates in these individuals. In absolute terms, this would imply as many as >2500 and >3800 colonoscopies in 50-year-old men and women, respectively, and still >500 and >800 colonoscopies in 70-year-old men and women, respectively.

**Figure 3.**
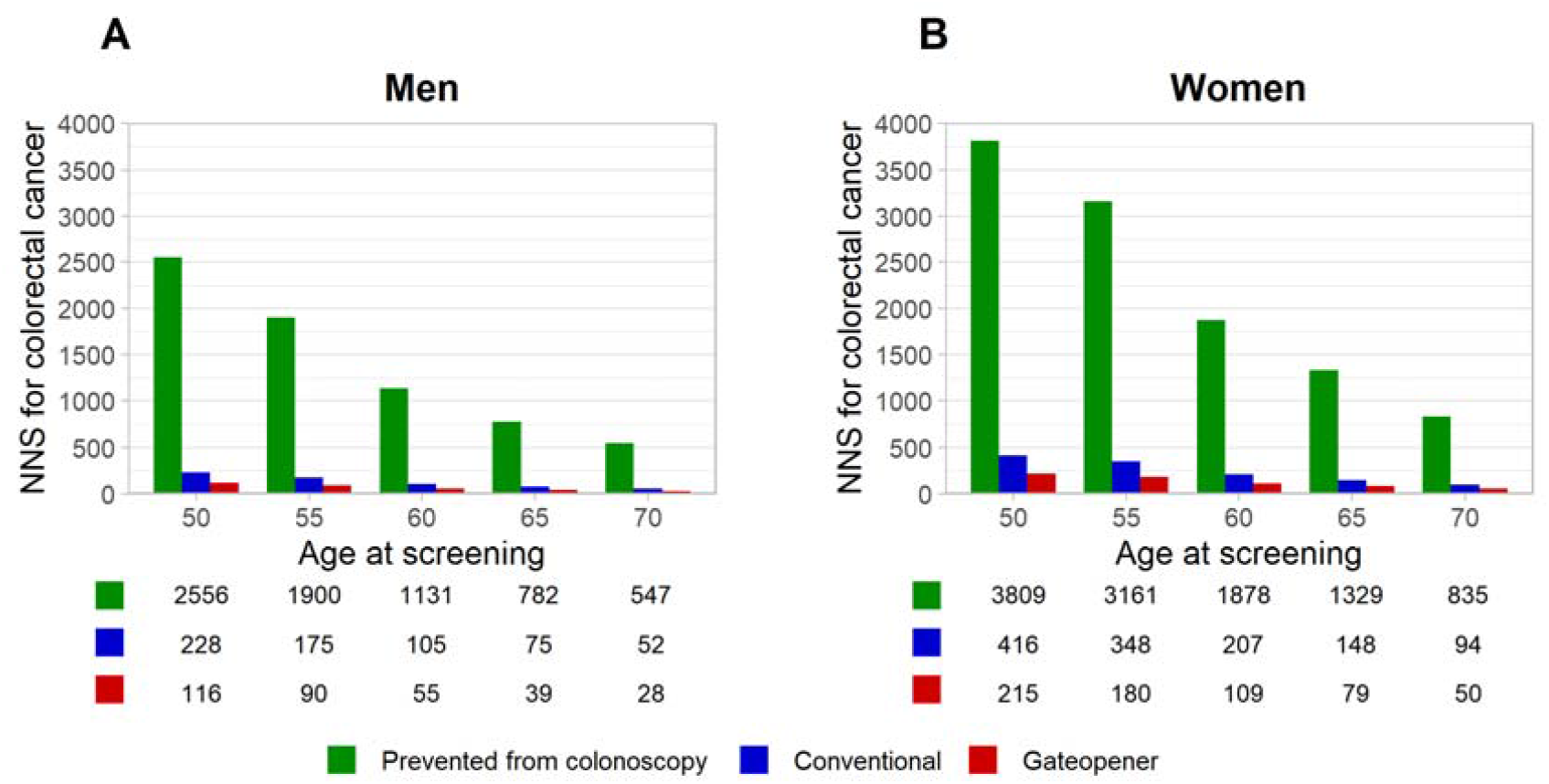
Number of colonoscopies needed to detect one case of colorectal cancer at varying ages of colonoscopy screening without and with positive preceding gateopener FIT, and in those in fact prevented from undergoing screening colonoscopy due to a negative preceding gateopener FIT. (A) men; (B) women. CRC: Colorectal cancer. NNS: number needed to scope.

Likewise, regardless of low, moderate, or high uptake of screening offers, the gateopener approach uniformly resulted in 51-53% more prevented CRC cases and 63-68% more prevented CRC deaths, respectively, within 10 years from colonoscopy screening. Detailed results according to sex and age are shown for moderate colonoscopy uptake rates (20%) in **Figure 4**, and for low (10%) and high (30%) uptake rates in **Supplementary Figures 3 and 4**, respectively. Relative gains in comparative effectiveness were consistent across all age groups and both sexes for prevented CRC cases and slightly increased along advancing age of screening for prevented CRC deaths.

**Figure 4.**
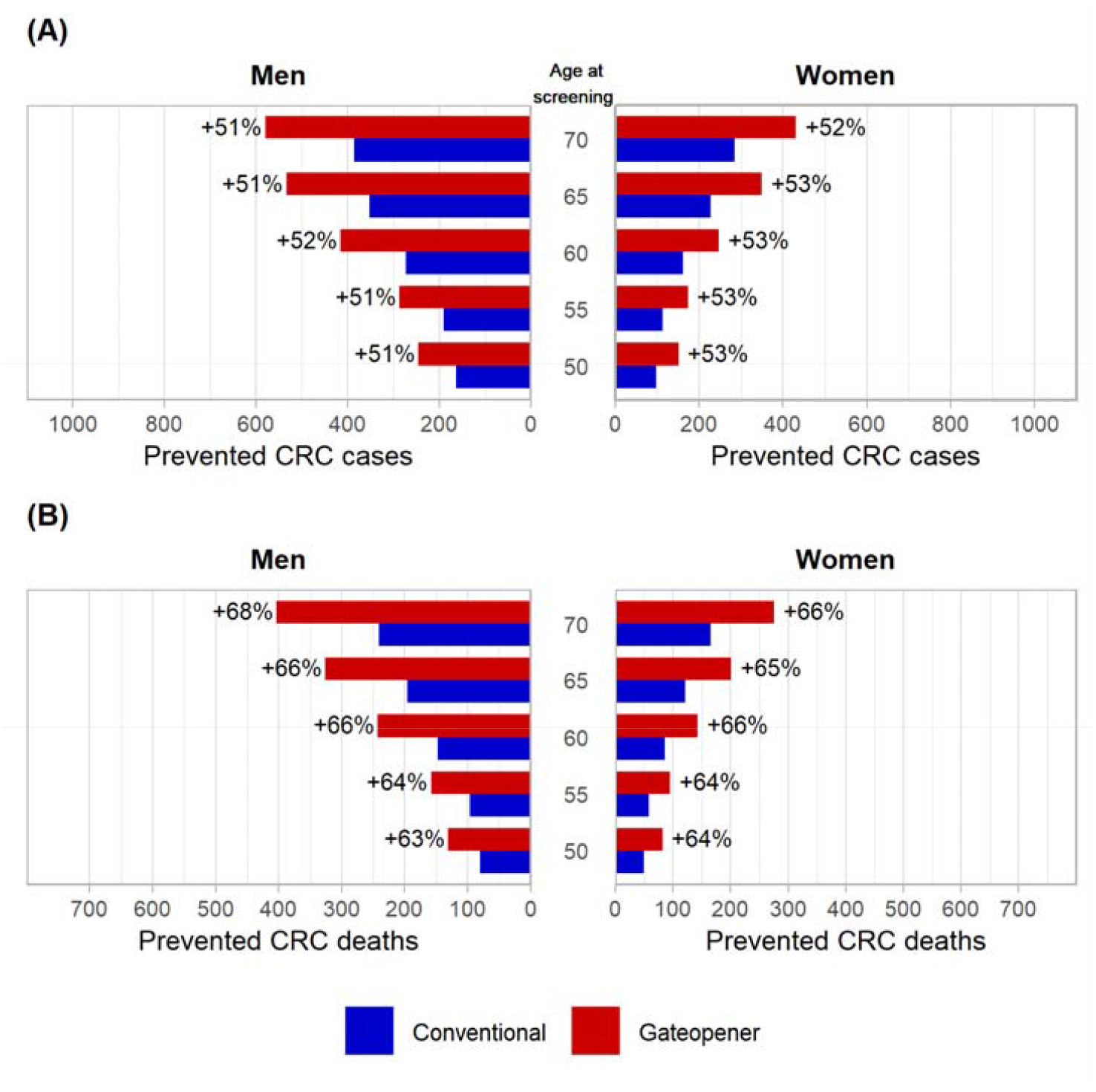
Expected numbers of prevented CRC cases and CRC deaths per 100,000 target population within 10 years after screening at identical, moderate levels of total colonoscopy use (20%) (A) Prevented CRC cases (B) Prevented CRC deaths CRC: Colorectal cancer, FIT: fecal immunochemical test

Gateopener scenarios with adherence levels resulting in 50% higher levels of overall colonoscopy use versus conventional screening (30% rather than 20%) more than doubled the numbers of prevented of CRC cases and deaths (**Figure 5)**. Sensitivity analyses using upper and lower limits of 95% CI of starting prevalences and annual transition rates yielded comparable patterns to the base case scenarios, and varying these parameters had overall very little impact on estimates on the relative effectiveness of gateopener vs conventional screening.

**Figure 5.**
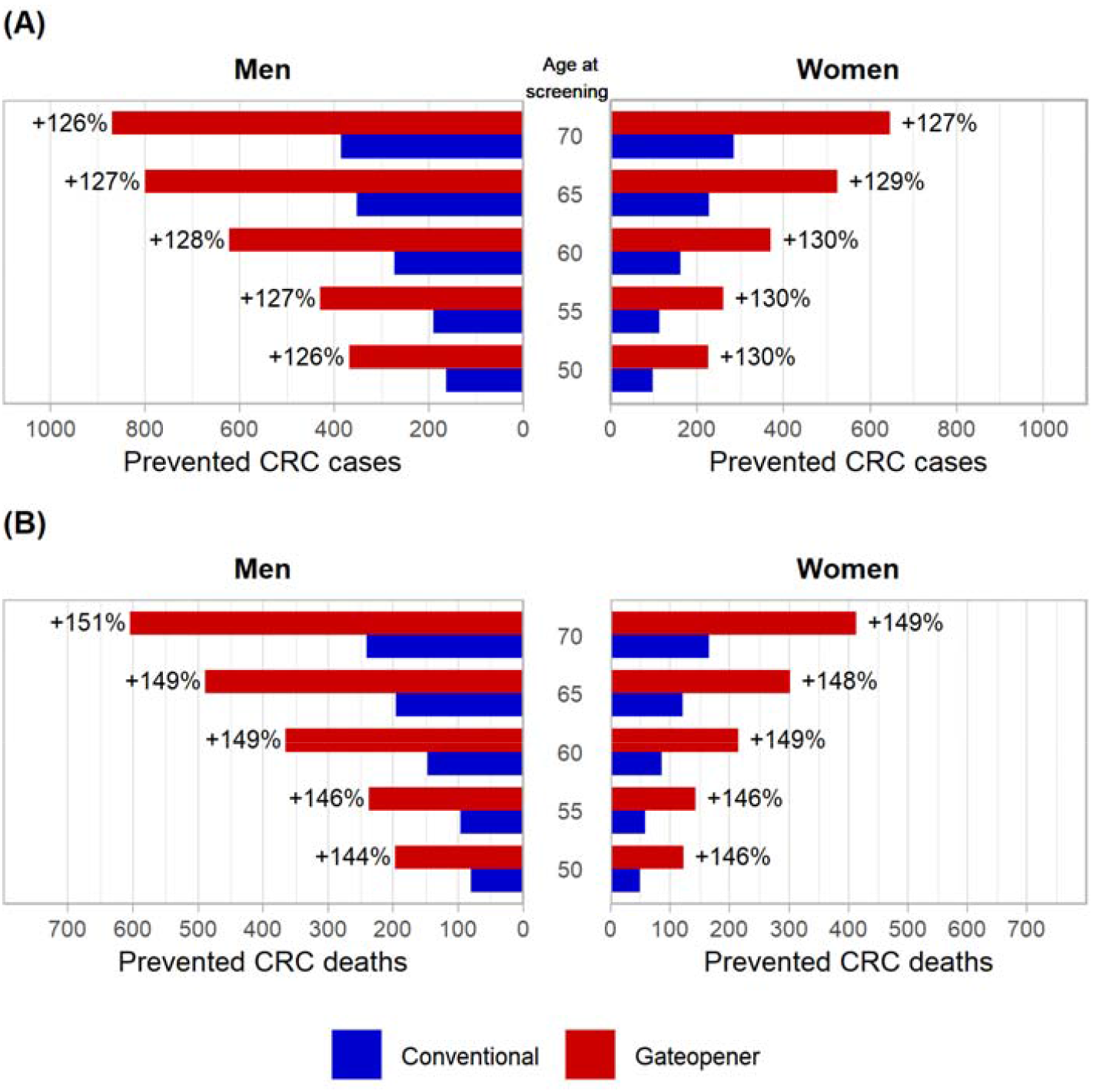
Expected numbers of prevented CRC cases and CRC deaths per 100,000 target population within 10 years after screening at varying levels of total colonoscopy use (gateopener screening, 30%, conventional screening colonoscopy, 20%) (A) Prevented CRC cases (B) Prevented CRC deaths CRC: Colorectal cancer, FIT: fecal immunochemical test

## Discussion

This simulation study assessed an innovative ‘gateopener’ approach to CRC screening in a hypothetical German population. When compared to conventional primary screening colonoscopy, the combination of a low threshold gateopener FIT followed by screening colonoscopy only offered to those with a positive stool test resulted in approximately doubled cancer detection rates, and thus 50% lower numbers of colonoscopies needed to detect one case of cancer, across both sexes and all assessed ages. The NNS in those spared from undergoing screening colonoscopy due to a negative gateopener FIT were approximately 10-times higher as compared to those after a positive FIT pretest, reflecting the low prevalence of colorectal neoplasms in those negatively pre-tested. Within ten years from screening, at identical levels of total colonoscopy use, the gateopener approach resulted in 51-53% and 63-68% more prevented CRC cases and deaths, respectively. Taking into account that gateopener screening may likely even result in higher levels of total colonoscopy use, corresponding scenarios with 50% higher colonoscopy uptake rates more than doubled the numbers of prevented CRC cases and deaths.

### Findings in context

Colonoscopy is generally considered the gold standard test for early detection of CRC and its precursors as it rarely misses advanced colonic lesions [22], and screening by this highly sensitive method has been estimated to reduce the risk of developing or dying from CRC by 60-70% [23]. A recent study showed that in countries with long-standing screening colonoscopy programs, CRC incidence and mortality decreased substantially over time [24]. Despite the large potential of colonoscopy-based screening, there are also major concerns related to the design and characteristics of conventional screening colonoscopy offers that may substantially limit the effectiveness of such offers and call for novel approaches to overcome them.

Firstly, as screening colonoscopy is typically offered in mostly unselected, average-risk populations, most screenees have no real benefit from this invasive procedure (which, albeit rarely, can also cause complications, such as bleedings [25]). Cancers and any advanced colorectal lesions are found in less than 1% and less than 10% of screening participants, respectively [7,26], as also reproduced by the conventional screening colonoscopy scenarios in this study. The vast majority of participants would never develop CRC even without undergoing this comparably elaborate procedure [21], which renders its use in unselected individuals rather inefficient.

Secondly, at the same time, conventional (primary) screening colonoscopy programs often fail to achieve adequate uptake by the eligible population [4]. This implies that those at high CRC risk are still being missed (for not using screening in the first place), even though most subjects undergoing the procedure will in fact not benefit. Substantially higher levels of population-wide adherence may be achieved in screening programs using a non-invasive test, such as a stool test, as primary screening test [5]. Well-organized programs, such as in the Netherlands, reported faecal test utilization rates of 60-80% [3]. By contrast, annual uptake rates of screening colonoscopy by the eligible population in Germany have been as low as 1-3 %, which translates to uptake rates within 10 years (the commonly recommended screening interval for colonoscopy) of 10-30% [27,28]. Even though uptake is also influenced by other characteristics of the screening program (such as invitation schemes) and the eligible population (such as age, sex, lifestyle, previous diagnostic colonoscopies, and cultural background [2]), such large differences may be attributed to a major extent to the low acceptability of screening colonoscopy as primary screening modality, mostly due to the discomfort associated with the procedure [29].

In this study, we proposed a gateopener approach which may be suited to address both described limiting factors in an innovative fashion, and thereby has great potential to considerably enhance the effectiveness of screening colonoscopy. Essentially, gateopener screening involves pre-selecting those subjects most likely to benefit, as only individuals above a low-level hemoglobin threshold of the gateopener FIT would be invited to proceed to screening colonoscopy. We demonstrated that such pre-selection would go along with a significantly higher diagnostic yield and CRC risk reductions at the same number of conducted colonoscopies as compared to conventional screening colonoscopy alone. A potential alternative application of the gateopener approach could be to enhance feasibility of screening colonoscopy in health systems with limited colonoscopy capacities. With a somewhat more restrictive positivity threshold, the number of screening colonoscopies could be substantially reduced while still maintaining comparable detection and prevention rates of colorectal neoplasms.

Furthermore, gateopener screening can be designed to optimally take advantage of factors known to be associated with increased uptake of screening. The required gateopener FIT as a means of pre-testing represents a low barrier to participation, as it is non-invasive and easy to use, does not require any dietary restrictions and only one sample is needed. Thus, uptake rates comparable to first rounds of conventional FIT screening appear plausible and were assumed for scenarios with moderate and high utilization levels calculated in this study [3]. One well-established measure to ensure adequate adherence is to directly include the FIT in mailed invitations for screening [20], thereby making its use straightforward and convenient. Further measures to increase uptake that could readily be combined include patient navigation, education measures and information campaigns [30]. We believe that these design features would likely imply higher colonoscopy utilization rates as compared to conventional screening colonoscopy. Combined with the inherently much more efficient use of colonoscopy capacities, this might readily even double the numbers of prevented CRC cases and deaths within ten years, as illustrated in a supplementary analysis.

Notably, a key conceptual strength of the gateopener approach is to make use of the potential of FITs to exactly quantify faecal haemoglobin concentrations, which has been largely ignored by now. FIT-based screening has continued to rely on screening strategies originally found effective for their predecessors, the traditional guaiac-based tests, i.e., at annual or biennial intervals and by using FITs in a qualitative rather than quantitative fashion. Subjects with a ‘positive’ result, with haemoglobin levels above a certain cut-off, are recommended to undergo colonoscopy for diagnostic work-up, while those with ‘negative’ results, i.e., levels below the cut-off, are re-invited for testing 1-2 years later. It should be noted that this differs fundamentally from the concept of gateopener screening, which would have screening colonoscopy at the typically recommended 10-yearly intervals still as its backbone, and a low-threshold FIT to pre-select those individuals most likely to benefit.

While there is some evidence on the use of lower cut-offs (and higher positivity rates) for FIT-based screening strategies, e.g. for defining FIT screening intervals [31–33] and of combined use of fecal occult blood testing and flexible sigmoidoscopy to enhance sensitivity of CRC screening [34–38], to our knowledge, no previous randomized, observational, or modelling study has assessed the performance of a screening strategy in which a screening colonoscopy-eligible population is pre-selected by use of a low-threshold, quantitative FIT.

### Strengths and Limitations

A major strength of our analysis is that key parameters used for the modeling by a thoroughly validated model were based on comprehensive analyses of data from a long-standing nationwide screening colonoscopy registry, the world’s largest of its kind, and detailed analyses of diagnostic value of FIT below commonly used thresholds to define FIT positivity from a large cohort of screening colonoscopy participants [17].

However, our study also has limitations that require careful consideration. Specific limitations of COSIMO have been described previously [8]. Briefly, major limitations concern model simplifying assumptions and uncertainties related to input parameters where evidence was limited. A major limitation of this study is that, due to the novelty and innovative character of the concept, no evidence on uptake patterns from real-life studies explicitly pre-selecting a colonoscopy-eligible population by a low-threshold FIT was available to inform our model. To account for this uncertainty, we simulated varying levels of adherence relying on previous evidence on FIT- and colonoscopy-based screening. However, the behavior of a population invited to gateopener screening is unknown. For instance, it is unclear whether offering a low-threshold FIT, which is associated with a high rate of false-positive test results, would impact adherence patterns. Real-life data, e.g., by a pilot study to also assess feasibility, would be needed to assess the plausibility of these assumptions.

Furthermore, this study is limited to considerations on clinical effectiveness. Further study from a health-economics point-of-view is warranted, as, for instance, the gateopener approach also requires considerable numbers of FIT test kits with associated additional costs. However, given that colonoscopy costs are multiple times higher than those for FIT test kits, we would not expect FIT pre-screening to offset the benefits of more targeted colonoscopy use to any relevant extent.

Finally, our analyses were limited to a 10-year time window following screening colonoscopy among previously unscreened people. Although once only colonoscopy has been suggested as a possible screening strategy, especially in populations with limited colonoscopy capacities, screening colonoscopy is mostly recommended to be repeated in 10-year intervals. Although a similar impact of the gateopener approach would be expected for repeat colonoscopies, we deliberately refrained from including repeat screening colonoscopy scenarios. The additional layers of complexity induced by various variants of “longitudinal adherence” [39–41] to various variants of repeat screening offers might have distracted from first-time demonstration of the key principles and implications of the gateopener approach. Further research should address a broader range of scenarios to more precisely derive expected gains of the gateopener approach in specific screening settings.

## Conclusion

In summary, our results suggest that the combination of a directly mailed, low-threshold FIT followed by screening colonoscopy might enable a considerably higher diagnostic yield and considerably stronger reduction of CRC incidence and mortality rates at a comparable “colonoscopy burden” than conventional screening colonoscopy strategies. By pre-selecting those most likely to benefit from it, such a ‘gateopener’ approach might be suited to substantially enhance the efficiency of colonoscopy-based CRC screening. Future studies assessing the potential of this concept in real-life screening practice are warranted.

## Data Availability

All analyses relevant to the study are included in the article or uploaded as supplementary information. The model source code is freely available from the DKFZ website (https://www.dkfz.de/de/klinepi/download/index.html).

