## Supplementary Material for "Making colonoscopy-based screening more efficient: a ‘gateopener’ approach"

Alternative Colonoscopy-Based Screening

Thomas Heisser, Rafael Cardoso, Tobias Niedermaier, Michael Hoffmeister,

Hermann Brenner

Table of Contents

**[SUPPLEMENTARY MATERIAL](#_Toc88140306)**	[1](#_Toc88140306)

### **Supplementary Appendix 1. COSIMO Model Documentation**

#### **Conceptual model structure**

Our Markov-based Colorectal Cancer Multistate Simulation Model (COSIMO) simulates the natural history of CRC based on the process of precursor lesions (non-advanced and advanced adenomas) developing into preclinical (asymptomatic) and then clinical (symptomatic) cancer. The simulation is performed on a hypothetical previously unscreened German population, with the number of simulated subjects and their corresponding baseline age (minimum 50 years) being variables to be chosen prior to model start. COSIMO can principally be used for simulating any population, provided updated or appropriately adjusted input parameters.

At start of the simulation, certain proportions of no neoplasm, non-advanced adenoma, advanced adenoma and preclinical CRC are assigned to the hypothetical population. The simulation runs up to a predefined number of cycles of each one year. Each year, people at each state have a certain probability (transition rate) to progress to the next state. Subjects with CRC may die from the disease, and at each state people may experience non-CRC death, reflecting the general background mortality from other causes.

Screening can alter the progression between states. People with adenoma will be moved backward to the state of no neoplasm, assuming removal of their adenoma at colonoscopy (for screening or diagnostic workup, e.g., after a positive fecal test). Subjects will then continue to have the probabilities to progress to the next states as those without findings at screening. We assume that, although these people are under a higher risk of developing adenomas or cancers than the general population [1], the excess risk will be effectively compensated through the protection provided by surveillance colonoscopies [2,3]. Preclinical CRC detected at screening will be moved forward to the state of diagnosed cancer.

After each cycle where a screening test was applied, the model differentiates the simulated population into a ‘screening negative’ and a ‘screening positive’ group, which allows for modelling different trajectories depending on the screening outcome. In such scenarios, subjects only receive the next screening round if they had a negative test result in the respective previous round. In the base case model, subjects with false-positive test results (e.g., a diagnostic colonoscopy without finding after a positive faecal immunochemical test) return to the screening population after a latency period of 10 years. Subjects with detected non-advanced adenomas are assumed to undergo surveillance colonoscopies at intervals of 10 years up to a predefined end age of 75 years. In case an advanced adenoma was detected, either at the primary screening test or at a surveillance colonoscopy, subjects are assumed to undergo periodic surveillance colonoscopies at three-yearly intervals up a predefined end age of 85.

The model source code, developed in the statistical software R (version 4.0.2), is available for download from our website [4].

#### **Model parameters**

**Starting prevalences and transition rates**

An overview of key model parameters is given in **Supplementary** **Table 1**.

*Data source*

The data basis of our analyses on model starting prevalences and transition rates was the nationwide screening colonoscopy registry run by the Central Research Institute of Ambulatory Health Care in Germany. The registry, which was built up along with the introduction of the screening colonoscopy offer in the year 2002, is a repository of all screening colonoscopies conducted in Germany. Reporting is virtually complete, as it is a prerequisite for physicians’ reimbursement by the health insurance funds. The registry includes only primary screening examinations (i.e., colonoscopies conducted for surveillance, work-up of symptoms or other screening tests are not included). Items reported include, besides basic sociodemographic variables, findings at colonoscopy, including number, size and histological characteristics of polyps. In case of multiple neoplasms, only the most advanced one (non-advanced adenoma, advanced adenoma, or cancer) is recorded. Advanced adenomas are defined as at least 1 adenoma ≥ 1 cm or at least 1 adenoma with villous components or high-grade dysplasia.

Noteworthy, the reporting for the screening colonoscopy registry does not differentiate by the class of lesion. Thus, the herein used term ‘adenoma’ refers to conventional or serrated adenomas (polyps) alike. While we preferred to refer to our model as being based on the adenoma-carcinoma pathway in previous publications [5–9] for the sake of simplicity and comprehensibility (as the grand majority of CRCs develops through this well-established pathway of cancer development [10,11]), in fact COSIMO’s defining parameters were derived using polyp/adenoma prevalences as detected and reported at screening colonoscopy, regardless of their underlying mechanism or pathway of development. Therefore, it will be more precise to refer to the model as being based on the ‘natural history of CRC’, without restrictions on underlying CRC development pathways.

*Starting prevalence*

The proportions of no neoplasm, non-advanced adenoma, advanced adenoma and preclinical CRC at the beginning of simulation were calculated based on the data from 344,658 participants of the German screening colonoscopy program who had their first screening colonoscopy during 2003–2012 at the age of 55 years [7]. To take into account that a certain proportion of neoplasms needs to be assumed to have been missed at colonoscopy screening, in particular for serrated or flat polyps [12,13], we re-calculated the previously reported prevalences [7], assuming representative miss rates of 25% for non-advanced adenomas and 5% for advanced neoplasms (advanced adenomas and preclinical cancers).

This was used as the best estimate for simulations starting with a 50-year-old population, which seems reasonable as selected regional programs which offer screening colonoscopy from age 50 on found similar prevalences of adenomas in age groups 50-54 and 55-59 [14].

*Transition rates*

Transition rates between states were estimated based on data from the nationwide screening colonoscopy registry by several separate birth cohort and mean sojourn time analysis. Details on the principles of these methods have been described previously [15–17]. Briefly, sex- and age-specific annual incidence and transition rates were estimated from sex- and age-specific prevalences of adenomas among 3.6 – 4.3 million screening participants from the same birth cohorts in 2003–2011 (2003-2009) and 2004–2012 (2004 – 2010). The analysis on mean sojourn time of preclinical cancers additionally incorporated registry-reported colorectal cancer incidence and participation rates in screening colonoscopy from 2003-2006.

Similar as for the starting prevalences, as colonoscopy was shown to be less effective in detecting serrated lesions (and as the true proportions of missed conventional adenomas and serrated lesions in the registry-reported prevalences is unknown), we re-calculated previously reported transition rates [15–17] to adjust for representative colonoscopy miss rates [12,13]. This adjustment resulted in slightly higher overall prevalences of adenomas, and therefore (when compared to previously reported rates) in slightly higher transition rates of incidence adenomas, as well as slightly lower transition rates from non-advanced to advanced adenomas and from adenomas to cancer. Furthermore, to adjust for uncertainties resulting from the cycle length of one year used in COSIMO, we updated the model to allow for small proportion of subjects with very rapidly progressing lesions with limited potential for early detection and associated worse prognosis. Age- and sex-specific annual transition rates between the states were estimated for age groups from 55-79 years in steps of 5 years. Estimates for age 50-54 and ≥ 80 (or ≥ 85) were assumed to be the same as those for age group 55-59 and 75-79 (or 80-84), respectively.

Confidence intervals for both starting prevalences and transition rates were derived by bootstrap analysis with resampling within sex- and age-specific subgroups. Ninety-five percent confidence intervals were determined as the 2.5th and 97.5th percentile of transition rate estimates obtained in 1,000 runs.

**Mortality rates**

Mortality rates for patients whose cancer was detected by screening or by symptoms were estimated in previous analyses [8,9]. We combined data on the proportion of screening-detected cases among all CRC cases in Germany during 2003-2012 in people aged 55-79 years [5,18] with the overall CRC-specific mortality rates by year after diagnosis in Germany in 2011-2012 [18]. We then used hazard ratios for patients detected by screening versus symptoms as obtained from a German population-based case-control study on CRC screening with long-term mortality follow-up of CRC patients [8,19] to estimate CRC-specific mortality rates by mode of detection (**Supplementary** **Table 2**). Sex- and age-specific general mortality rates and average life expectancy of the population were extracted from German population life tables 2010/2012 (**Supplementary** **Table 3)** [20].

**Model validation**

COSIMO has been validated for the German screening-eligible population. Details on the model validation process can be found in the literature [21]. Briefly, we pursued a three-fold approach using the best available evidence from epidemiological data sources in Germany. We compared model-derived cumulative incidence and prevalences of colorectal neoplasms to (a) results from KolosSal, a study in German screening colonoscopy participants, (b) registry-based estimates of CRC incidence in Germany, and (c) outcome patterns of randomized sigmoidoscopy screening studies. This approach enabled us to scrutinize the model's natural history component (Parts a and b) as well as the modeled effect of screening colonoscopy (Parts b and c) at the same time.

We found that (a) more than 90% of observed prevalences in the KolosSal study were within the 95% confidence intervals of the model-predicted neoplasm prevalences; (b) the 15-year cumulative CRC incidences estimated by simulations for the German population deviated by 0.0% to 0.2% units in men and 0.0% to 0.3% units in women when compared to corresponding registry-derived estimates; and (c) the time course of cumulative CRC incidence and mortality in the modeled intervention group and control group closely resembles the time course reported from sigmoidoscopy screening trials. Overall, COSIMO adequately predicted colorectal neoplasm prevalences and incidences in a German population for up to 25 years, with estimated patterns of the effect of screening colonoscopy resembling those seen in registry data and real-world studies.

### **Supplementary Appendix 2. Supplementary Tables and Figures**

**Overview**

**Supplementary Table 1.** Overview of model parameters

**Supplementary Table 2.** Annual CRC-specific mortality rates of CRC patients by mode of cancer detection

**Supplementary Table 3.** Sex- and age-specific general mortality rates

**Supplementary Figure 1**. Detection rates of any advanced neoplasm (advanced adenoma or cancer) at varying ages of colonoscopy screening without and with positive preceding ‘gateopener’ FIT, and in those in fact prevented from undergoing screening colonoscopy due to a negative preceding ‘gateopener’ FIT

**Supplementary Figure 2**. Number of colonoscopies needed to detect one advanced neoplasm (advanced adenoma or cancer) at varying ages of colonoscopy screening without and with positive preceding ‘gateopener’ FIT, and in those in fact prevented from undergoing screening colonoscopy due to a negative preceding ‘gateopener’ FIT

**Supplementary Figure 3.** Expected numbers of prevented CRC cases and CRC deaths per 100,000 target population within 10 years after screening at identical levels of total colonoscopy use (10%)

**Supplementary Figure 4.** Expected numbers of prevented CRC cases and CRC deaths per 100,000 target population within 10 years after screening at identical levels of total colonoscopy use (30%)

#### **Supplementary Table 1.** Overview of model parameters

| **A. Proportions of no neoplasm, non-advanced adenoma, advanced adenoma and preclinical CRC at the beginning of simulation^1^** | | | | | | | | | |
| --- | --- | --- | --- | --- | --- | --- | --- | --- | --- |
|  |  | **Most advanced finding**  **% (95% confidence interval)** | | | | | | | |
| **Sex** |  | **No neoplasm** | | **Non-advanced adenoma** | | **Advanced adenoma** | | **Preclinical colorectal cancer** | |
| Men |  | 71.5 (71.3 – 71.7) | | 21.7 (21.5 – 21.9) | | 6.3 (6.1 – 6.4) | | 0.48 (0.45 – 0.52) | |
| Women |  | 83.2 (83.0 – 83.3) | | 13.2 (13.0 – 13.3) | | 3.4 (3.3 – 3.5) | | 0.26 (0.24 – 0.29) | |
| ^1^ Estimates based on the German screening colonoscopy registry. Extracted and recalculated from reference [7] | | | | | | | | | |
| **B. Sex- and age-specific annual transition rates between states²** | | | | | | | | |  |
|  |  | **Annual transition rates**  **% (95% confidence interval)** | | | | | | |  |
| **Sex** | **Age** | **No neoplasm to non-advanced adenoma** | **Non-advanced adenoma to advanced adenoma** | | **Advanced adenoma to preclinical colorectal cancer** | | **Preclinical colorectal cancer to clinical colorectal cancer** | | **Preclinical colorectal cancer to colorectal cancer death** |
| Men | 50-54 | 3.1 (2.9 – 3.4) | 3.3 (2.8 – 3.9) | | 2.6 (2.2 – 3.1) | | 15.5 (14.9 – 16.6) | | 1.5 (1.4 – 1.6) |
|  | 55-59 | 3.1 (2.9 – 3.4) | 3.3 (2.8 – 3.9) | | 2.6 (2.2 – 3.1) | | 15.5 (14.9 – 16.6) | | 1.5 (1.4 – 1.6) |
|  | 60-64 | 3.1 (2.8 – 3.4) | 3.2 (2.6 – 3.7) | | 3.1 (2.6 – 3.4) | | 16.4 (15.7 – 17.4) | | 1.6 (1.6 – 1.7) |
|  | 65-69 | 3.2 (2.9 – 3.4) | 3.2 (2.6 – 3.7) | | 3.8 (3.4 – 4.3) | | 18.2 (17.4 – 19.1) | | 1.8 (1.7 – 1.9) |
|  | 70-74 | 2.9 (2.6 – 3.3) | 3.3 (2.6 – 4.0) | | 5.1 (4.5 – 5.8) | | 17.6 (16.8 – 18.5) | | 1.7 (1.7 – 1.8) |
|  | 75-79 | 2.3 (1.8 – 2.9) | 3.0 (1.9 – 4.2) | | 5.2 (4.2 – 6.2) | | 17.3 (16.3 – 18.3) | | 1.7 (1.6 – 1.8) |
|  | 80+ | 2.3 (1.8 – 2.9) | 3.0 (1.9 – 4.2) | | 5.2 (4.2 – 6.2) | | 15.7 (14.5 – 17.1) | | 1.6 (1.4 –1.7) |
| Women | 50-54 | 1.8 (1.7 – 2.0) | 3.2 (2.6 – 3.8) | | 2.5 (2.0 – 2.9) | | 18.2 (16.8 – 19.7) | | 1.9 (1.8 – 2.1) |
|  | 55-59 | 1.8 (1.7 – 2.0) | 3.2 (2.6 – 3.8) | | 2.5 (2.0 – 2.9) | | 18.2 (16.8 – 19.7) | | 1.9 (1.8 – 2.1) |
|  | 60-64 | 2.0 (1.8 – 2.2) | 2.9 (2.2 – 3.4) | | 2.7 (2.2 – 3.2) | | 19.1 (17.8 – 20.3) | | 2.0 (1.9 – 2.1) |
|  | 65-69 | 2.1 (1.9 – 2.3) | 2.9 (2.3 – 3.5) | | 3.8 (3.3 – 4.3) | | 18.7 (17.7 – 19.7) | | 2.0 (1.9 – 2.1) |
|  | 70-74 | 2.0 (1.7 – 2.2) | 3.8 (3.0 – 4.6) | | 5.0 (4.2 – 5.7) | | 17.8 (16.8 – 18.9) | | 1.9 (1.8 – 2.0) |
|  | 75-79 | 1.6 (1.1 – 2.0) | 3.0 (1.7 – 4.4) | | 5.6 (4.4 – 6.8) | | 16.5 (15.5 – 17.7) | | 1.7 (1.6 – 1.9) |
|  | 80+ | 1.6 (1.1 – 2.0) | 3.0 (1.7 – 4.4) | | 5.6 (4.4 – 6.8) | | 14.9 (13.9 – 16.1) | | 1.6 (1.4 – 1.7) |
| ² Estimates extracted and recalculated from references [15–17] | | | | | | | | |  |

**Supplementary Table 1** Overview of model parameters (continued)

| **C. Diagnostic performance parameters** | | | | | |
| --- | --- | --- | --- | --- | --- |
|  |  | **Performance (%)** | | | |
| **Test (sex)** | **Parameter** | **No neoplasm** | **Non-advanced adenoma** | **Advanced adenoma** | **Preclinical colorectal cancer** |
| FIT (men) **³** | Sensitivity | - | 49.8 | 70.5 | 95.3 |
|  | Specificity | 54.0 | - | - | - |
| FIT (women) **³** | Sensitivity | - | 51.0 | 71.1 | 94.4 |
|  | Specificity | 52.3 | - | - | - |
| Colonoscopy (both sexes) ^4^ | Sensitivity | - | 75.0 | 95.0 | 95.0 |
|  | Specificity | 100 | - | - | - |
| ^3^ Sentinel FOBT Gold adjusted for a positivity rate of 50%. Corresponding cutoffs (µg/g): Men, 4.6. Women, 3.6  ^4^ Estimates based on references [12,13]  FIT, fecal immunochemical test | | | | | |

#### **Supplementary Table 2.** Annual CRC-specific mortality rates of CRC patients by mode of cancer detection^1^

|  | **Annual CRC-specific mortality rates (%)** | | | |
| --- | --- | --- | --- | --- |
| **Year after diagnosis** | **Screening colonoscopy– detected cases** | | **Symptom-detected cases** | |
|  | **Men** | **Women** | **Men** | **Women** |
| 1 | 4.6 | 3.7 | 19.7 | 20.6 |
| 2 | 2.2 | 1.9 | 9.3 | 10.7 |
| 3 | 2.1 | 1.3 | 8.8 | 7.4 |
| 4 | 1.5 | 0.9 | 6.3 | 4.8 |
| 5 | 1.2 | 0.6 | 5.0 | 3.3 |
| 6 | 0.8 | 0.3 | 3.5 | 1.7 |
| 7 | 0.4 | 0.3 | 1.8 | 1.8 |
| 8 | 0.4 | 0.3 | 1.9 | 1.8 |
| 9 | 0.4 | 0.0 | 1.9 | 0.0 |
| 10 | 0.0 | 0.0 | 0.0 | 0.0 |

^1^ Estimates extracted from references [8,9]

CRC, Colorectal cancer.

#### **Supplementary Table 3.** Sex- and age-specific general mortality rates

|  | **General mortality rates from age to age +1 (%)**^1^ | |
| --- | --- | --- |
| **Age** | **Men** | **Women** |
| 50 | 0.4 | 0.2 |
| 51 | 0.4 | 0.2 |
| 52 | 0.5 | 0.3 |
| 53 | 0.6 | 0.3 |
| 54 | 0.6 | 0.3 |
| 55 | 0.7 | 0.4 |
| 56 | 0.7 | 0.4 |
| 57 | 0.8 | 0.4 |
| 58 | 0.9 | 0.4 |
| 59 | 1.0 | 0.5 |
| 60 | 1.0 | 0.5 |
| 61 | 1.1 | 0.6 |
| 62 | 1.2 | 0.6 |
| 63 | 1.3 | 0.7 |
| 64 | 1.4 | 0.7 |
| 65 | 1.5 | 0.8 |
| 66 | 1.7 | 0.9 |
| 67 | 1.8 | 0.9 |
| 68 | 1.9 | 1.0 |
| 69 | 2.1 | 1.1 |
| 70 | 2.2 | 1.2 |
| 71 | 2.4 | 1.3 |
| 72 | 2.7 | 1.4 |
| 73 | 3.0 | 1.6 |
| 74 | 3.3 | 1.8 |
| 75 | 3.7 | 2.1 |
| 76 | 4.1 | 2.4 |
| 77 | 4.6 | 2.7 |
| 78 | 5.2 | 3.1 |
| 79 | 5.8 | 3.6 |

*Continued on next page*

***Supplementary* Table 3.** *Sex- and age-specific general mortality rates (continued)*

|  | **General mortality rates from age to age +1 (%)**^1^ | |
| --- | --- | --- |
| **Age** | **Men** | **Women** |
| 80 | 6.5 | 4.1 |
| 81 | 7.2 | 4.7 |
| 82 | 8.0 | 5.4 |
| 83 | 8.9 | 6.2 |
| 84 | 9.9 | 7.1 |
| 85 | 11.1 | 8.2 |
| 86 | 12.3 | 9.3 |
| 87 | 13.7 | 10.7 |
| 88 | 15.3 | 12.1 |
| 89 | 16.9 | 13.7 |
| 90 | 18.7 | 15.4 |
| 91 | 20.7 | 17.2 |
| 92 | 22.7 | 19.1 |
| 93 | 24.8 | 21.1 |
| 94 | 27.0 | 23.2 |
| 95 | 29.1 | 25.3 |
| 96 | 31.2 | 27.4 |
| 97 | 33.2 | 29.6 |
| 98 | 35.1 | 31.7 |
| 99 | 37.2 | 34.0 |
| 100 | 39.2 | 36.2 |

^1^Estimates were extracted from German population life tables 2010/2012 (reference [20]).

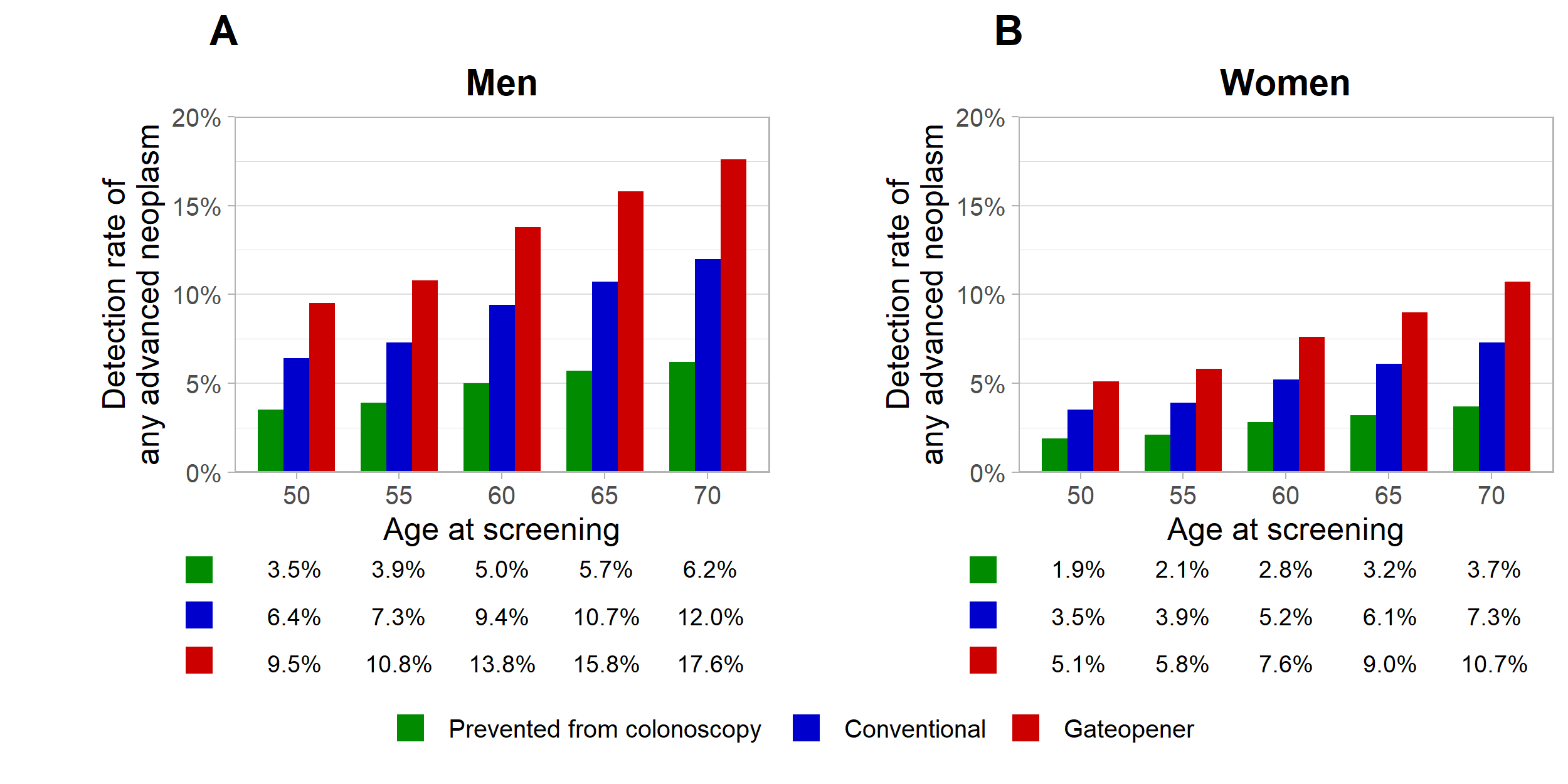

#### **Supplementary Figure 1.** Detection rates of any advanced neoplasm (advanced adenoma or cancer) at varying ages of colonoscopy screening without and with positive preceding ‘gateopener’ FIT, and in those in fact prevented from undergoing screening colonoscopy due to a negative preceding ‘gateopener’ FIT

(A) men; (B) women.

CRC: Colorectal cancer.

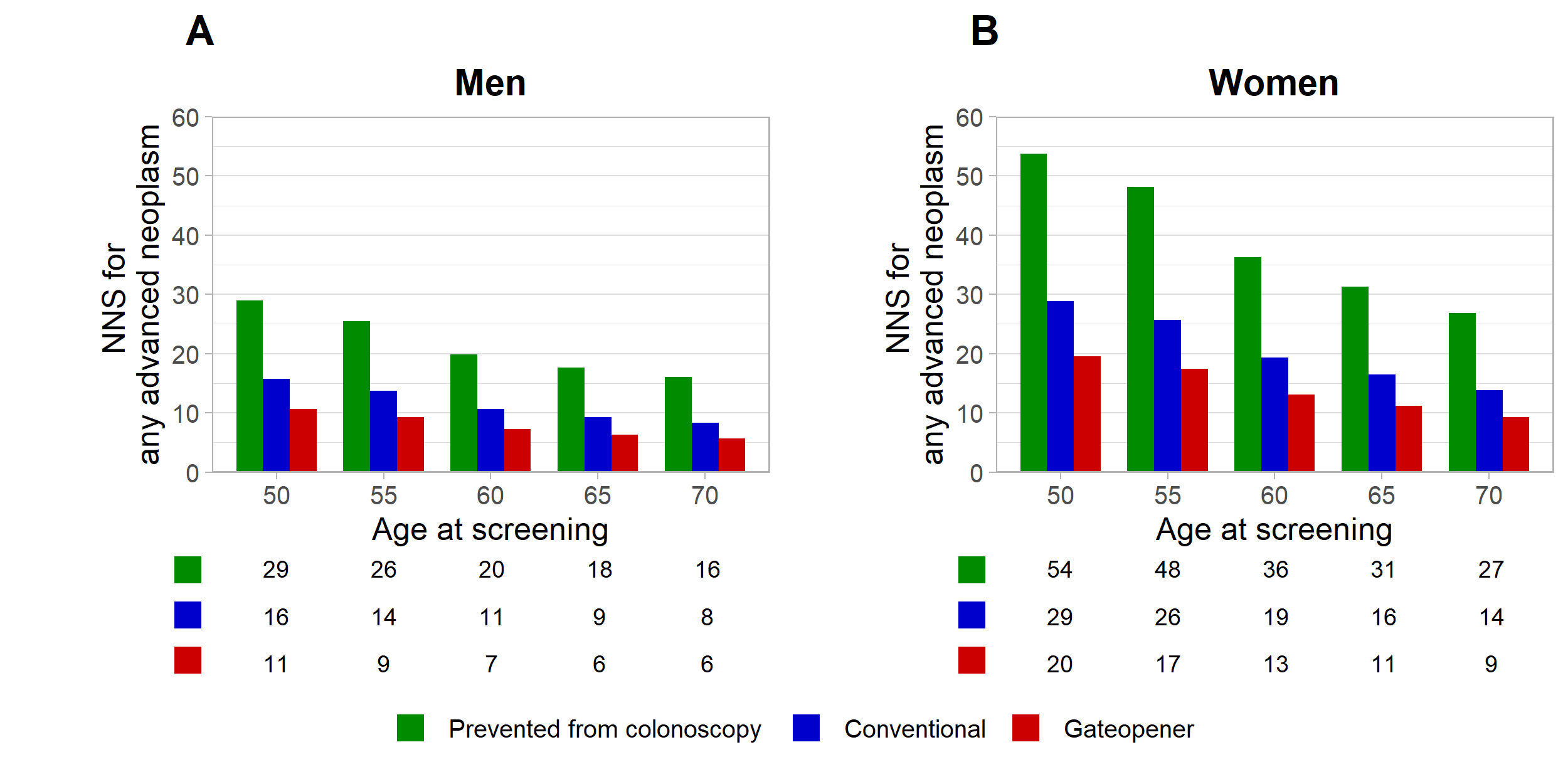

#### **Supplementary Figure 2.** Number of colonoscopies needed to detect one advanced neoplasm (advanced adenoma or cancer) at varying ages of colonoscopy screening without and with positive preceding ‘gateopener’ FIT, and in those in fact prevented from undergoing screening colonoscopy due to a negative preceding ‘gateopener’ FIT

(A) men; (B) women.

CRC: Colorectal cancer.

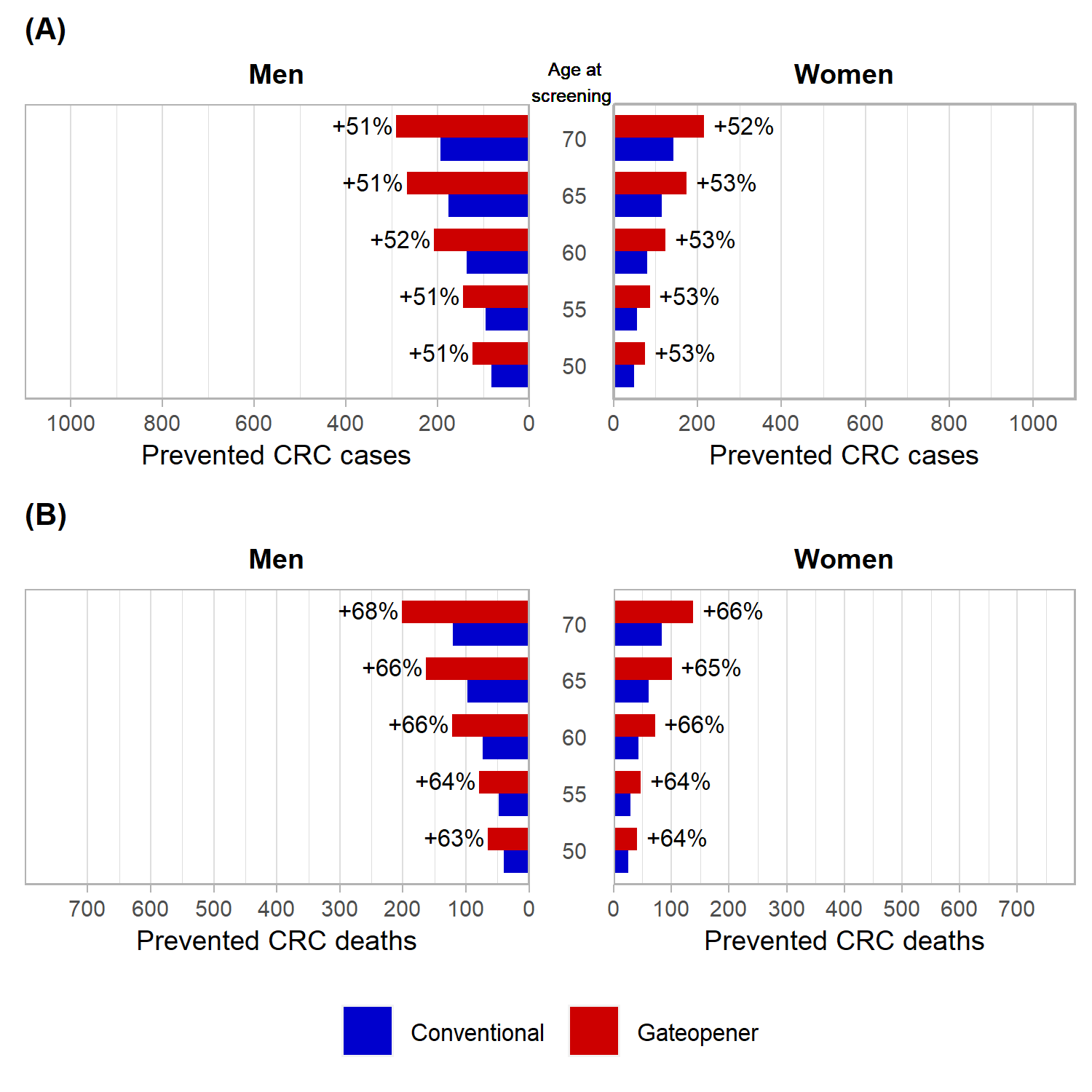

#### **Supplementary Figure 3.** Expected numbers of prevented CRC cases and CRC deaths per 100,000 target population within 10 years after screening at identical levels of total colonoscopy use (10%)

(A) Prevented CRC cases

(B) Prevented CRC deaths

CRC: Colorectal cancer, FIT: fecal immunochemical test

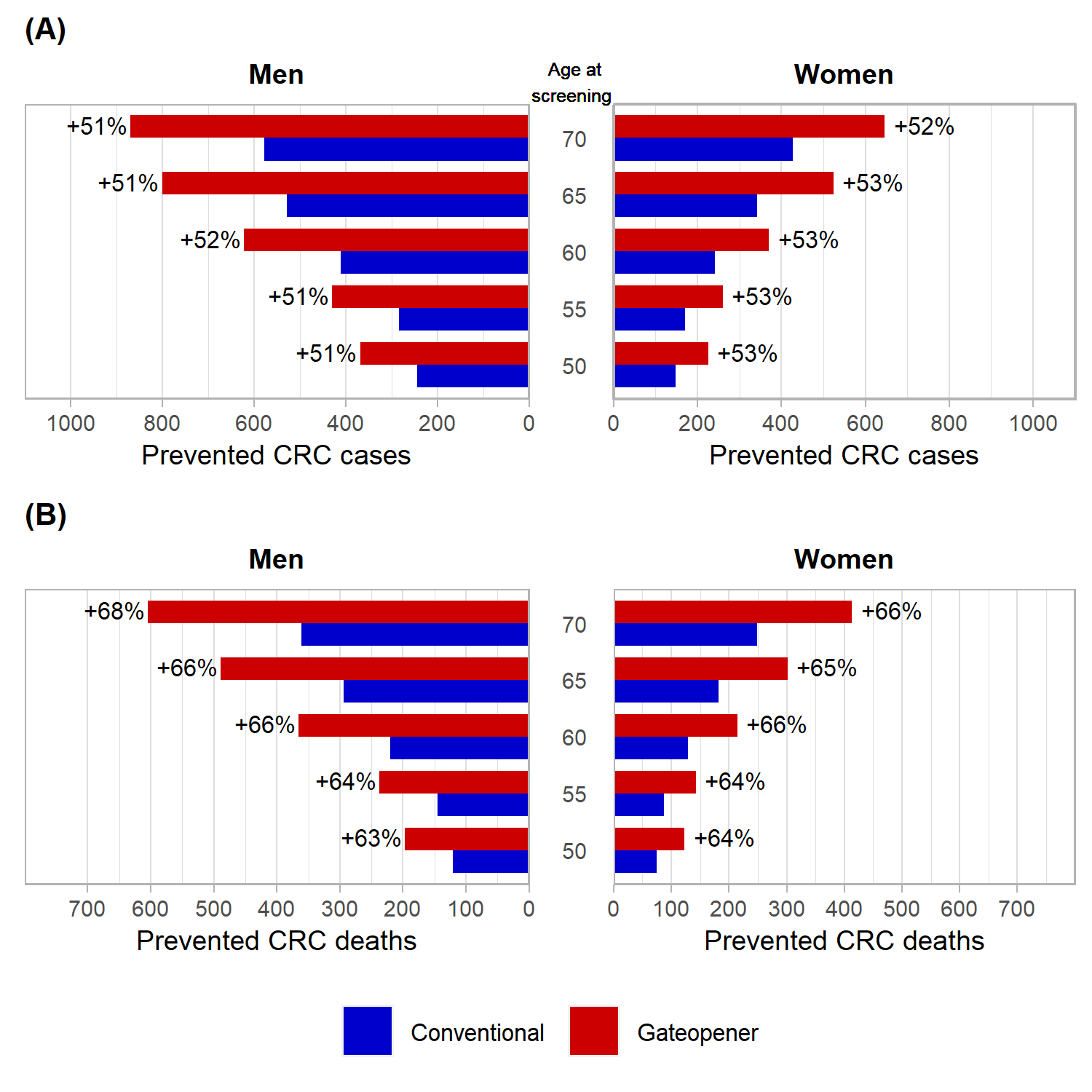

#### **Supplementary Figure 4.** Expected numbers of prevented CRC cases and CRC deaths per 100,000 target population within 10 years after screening at identical levels of total colonoscopy use (30%)

(A) Prevented CRC cases

(B) Prevented CRC deaths

CRC: Colorectal cancer, FIT: fecal immunochemical test
